# ATLAS: An automated association test using probabilistically linked health records with application to genetic studies

**DOI:** 10.1101/2021.05.02.21256490

**Authors:** Harrison G. Zhang, Boris P. Hejblum, Griffin M. Weber, Nathan P. Palmer, Susanne E. Churchill, Peter Szolovits, Shawn N. Murphy, Katherine P. Liao, Isaac S. Kohane, Tianxi Cai

## Abstract

**Objective:** Large amounts of health data are becoming available for biomedical research. Synthesizing information across databases with no gold standard mappings between records may provide a more complete picture of patient health and enable novel research studies. To do so, researchers may probabilistically link databases and conduct inference using the linked data. However, previous inference methods for linked data are constrained to specific linkage settings and exhibit low power. Here, we present ATLAS, an automated, flexible, and robust association testing algorithm for probabilistically linked data.

**Materials and Methods:** Missing variables are imputed at various thresholds using a weighted average method that propagates uncertainty from the linkage process. Next, an estimated effect size is obtained using a generalized linear model. ATLAS then conducts the threshold combination test by optimally combining p-values obtained from data imputed at varying thresholds using Fisher’s method and perturbation resampling.

**Results:** In simulations, ATLAS controls for type I error and exhibits high power compared to previous methods. In a real-world application study, incorporation of linked data-enabled analyses using ATLAS yielded two additional signifigant associations between rheumatoid arthritis genetic risk score and biomarkers.

**Discussion:** The ATLAS weighted average imputation weathers false matches and increases contribution of true matches to mitigate linkage error induced bias. ATLAS’ threshold combination test avoids arbitrarily choosing a threshold to rule a match, thus automating linked data-enabled analyses and preserving power.

**Conclusion:** ATLAS promises to enable novel and powerful research studies using linked data to capitalize on all available data sources.

## 1. Introduction

A vast amount of health data stemming from electronic health records (EHR), biorepositories, administrative claims, and biomedical research studies are becoming available for discovery and predictive research [1,2]. For patients who contribute information to multiple databases, synthesizing their information across all available sources captures a more complete picture of their health and allows for more comprehensive and powerful research studies. For example, database A may contain genomic data and database B may contain longitudinal phenotypic data. Linking patient records in these databases would allow researchers to investigate gene-disease associations. In a similar vein, researchers recently linked genomics data with environmental factors of BRCA1/BRCA2 mutation carriers from two independent studies to study environmental-gene relations, demonstrating the potential to conduct innovative research studies after linking databases [3].

To perform linkage when protected health information (PHI) identifiers are not available, as is generally the case in de-identified research databases, researchers employ probabilistic record linkage (PRL) [4]. However, linkage errors are inevitable in PRL due to data discrepancies, and examples of linkage errors include incorrectly linking two records that do not belong to the same patient (false matches) or leaving a record unlinked when a correct link exists (missed matches). Neter *et al*. were the first to investigate the consequences of linkage errors on downstream inference results, and they showed that such errors induce substantial bias in inference [5]. More recently, Rentsch *et al*. attempted to perform inference on linked real-world data and provided evidence that false matches reduce magnitudes of association while leading to highly biased estimates [6]. We are specifically interested in testing the association between some predictors *X* – recorded in one database A, and an outcome *Y* – recorded in a different database B. Within an association testing framework, both false matches and missed matches drive the results in the direction of no association by undermining statistical power [7–9]. This is because false matches increase sample sizes but dilute potential associations while missed matches reduce sample sizes and undermine statistical efficiency [7–9].

Despite the rapid growth in analysis of linked data and the well documented effects of linkage error on downstream inference, few robust, automated, and flexible inference methods have been proposed to account for linkage error induced bias [10]. Further, current proposed estimators are restricted to specific linkage settings and few are implemented in open-source software. For example, Hof *et al*. propose to address uncertainty from PRL by weighting least square estimators in linear and logistic regression [11]. But, their model does not formally account for non-match events, and they conclude that their estimator is biased unless complete matching is achieved [11]. Chipperfield proposes a weighting approach to make inference about regression coefficients but assumes that database *B* must be a complete subset of database *A* [12]. Dalzell *et al*.’s inference method necessitates the selection of extraneous blocking variables and, they demonstrate that block structure is needed to make unbiased estimates [13]. Recently, Han *et al*. propose linkage bias correcting estimators that do not make specific assumptions about linkage settings or require specific data structures as previously proposed methods do [14]. However, their method does not account for linkage settings where covariates come from either database *A* or *B* [14].

In this article, we propose automated association testing using probabilistically linked health records (ATLAS), a fully automated and scalable association testing framework that addresses many of these limitations. ATLAS utilizes either (1) a best match method or (2) a weighted imputation method that propagates uncertainty from the linkage process contained in matching probabilities. Then, ATLAS optimally combines several p-values estimated using generalized linear models (GLMs) that each correspond to a different matching threshold *ρ*_*k*_ (*k* ∈ {1, …, *K*}) as a significance test, avoiding the difficult choice of choosing a single threshold for defining a match. Unlike previous work, ATLAS performance is not conditional on specific linkage settings, linkage methods, or data structure. To facilitate ease of use and accessibility, we have implemented ATLAS as an open-source R package on CRAN. Here, we validate ATLAS performance and compare to existing methods in simulation and real-world studies to show that ATLAS is more robust at detecting associations than previously published estimators.

## 2. Materials and Methods

### 2.1. Statistical method

The proposed ATLAS algorithm broadly consists of 3 steps: (1) missing variable imputation using either i) the best match above a pre-specified threshold, or ii) a weighted average using matching probabilities as weights also filtering on a pre-specified threshold; (2) estimation of an adjusted effect size using a GLM; and (3) a significance test relying on optimal combination of p-values obtained from data imputed at varying thresholds using Fisher’s method together with perturbation resampling. Figure 1 illustrates this procedure and each of these three steps is further detailed in following subsections. Without loss of generality and for the sake of simplicity, here we will consider the situation where we have database A containing a *p*-dimensional novel predictor vector *X* and a vector of matching features *M*_*A*_ on *n*_*A*_ subjects indexed by *i* ∈ {1, …, *n*_*A*_} and database B containing outcome information *Y*, a vector of matching features *M*_*B*_, and potentially some other existing covariates *W* on *n*_*B*_ subjects indexed by *j* ∈ {1, …, *n*_*B*_}. We seek to link the predictor variables *X* recorded in database A subjects in database B such that we may run an association analysis of *Y* ∼ *W + X*.

**Figure 1:**
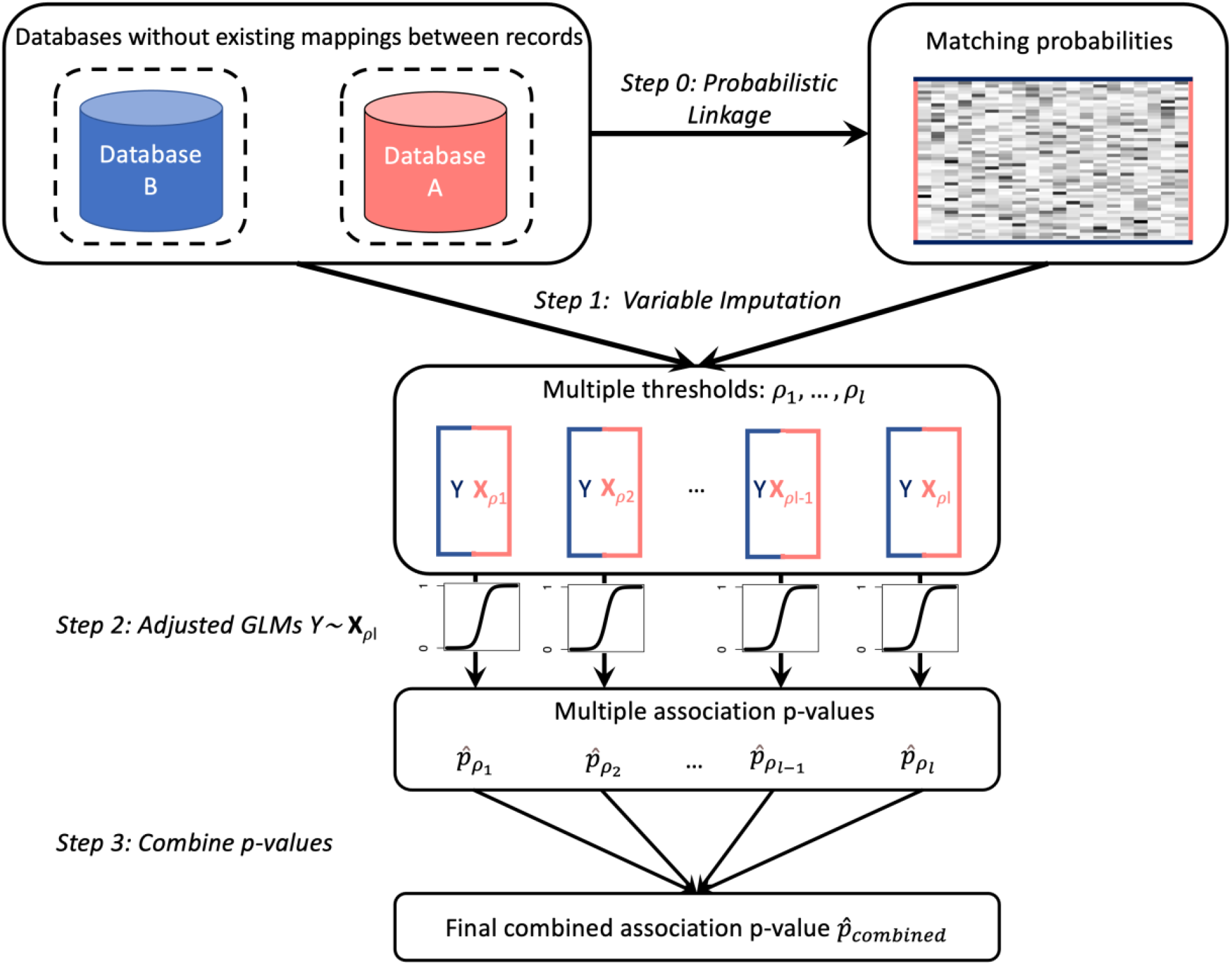
Schematic of the proposed ATLAS algorithm

#### 2.1.1. Missing variable imputation

The linkage problem can be seen as a missing information problem. We denote the probability of matching between a patient *i* from database *A* and a patient *j* from database *B* as π_*ij*_, where π_*ij*_ is ascertained via any given linkage algorithm that typically assess the similarity between the matching features *M*_*Ai*_ and *M*_*Bj*_. For example, the `ludic` linkage algorithm employs Bayesian modeling of binary diagnosis codes as matching features to estimate a posterior probability of being a match given a pair of patient records [15]. Using these estimated probabilities and given a specific matching cutoff threshold *ρ*_*k*_ above which patients are ruled as a match, we define *ξ*_*j*_(*ρ*) as whether there is any match for patient *j* in database B:

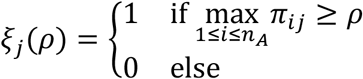

The best match imputation method imputes missing a *Z*_*j*_ with the observation *X*_*i*_ from patient *i* in *A* with the highest probability 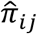 above a threshold *ρ*_*k*_. However, this method risks imputing missing variables from false matches created by linkage errors and diluting potential associations. Therefore, we propose instead a weighted average imputation of missing variables using linkage probabilities as weights to weather false matches and increase contribution of true matches. More specifically, for the *j*^*th*^ subject in database B and a given threshold *ρ*, we identify subjects in database A with linkage probabilities 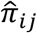 above *ρ* and obtain a weighted average of their *X*’s as the linked predictor:

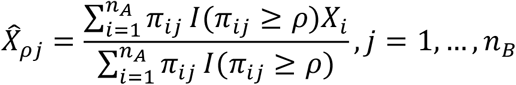

If 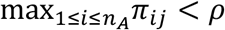, then the predictor will be deemed as missing for the *j*^*th*^ subject.

#### 2.1.2. Effect size estimation using Generalized Linear Models (GLM)

Our goal is to assess the association between *X* and *Y* using the linked database B through a GLM with link function *g*:

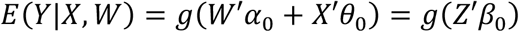

where *Z* = (*W′, X′*)*′* and *β* = (*α ′, θ* ′)*′*. For simplicity, we focus on the downstream association testing for *H*_*0*_: *θ* = 0 using linked data

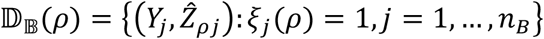

where 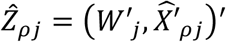. We fit the GLM 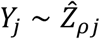 to 𝔻_𝔹_ to obtain a maximum likelihood estimate for *β*, denoted as 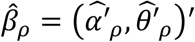, and test for *H*_*0*_: *θ*_0_= 0 based on the corresponding 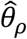.

#### 2.1.3. The ATLAS threshold combination test: significance testing with optimal p-value combination

Various threshold values can be used for *ρ*. For instance, one could use *ρ* = 0.5, where indicated matches have a higher probability of being match than non-match, or *ρ* = 0.9, where indicated matches have higher certainty of being true matches. However, the optimal choice of such a threshold is unclear in practice due to the lack of gold standard labels on the true mappings between A and B. On the one hand, higher thresholds have lower estimation biases from fewer false matches but at the price of decreased statistical power from smaller sample sizes. On the other hand, lower thresholds exhibit higher statistical power at the expense of increased estimation bias.

Thus, instead of arbitrarily choosing a threshold *ρ*, we propose to optimally combine several p-values that correspond to different matching thresholds {*ρ*_*ℓ*_, *ℓ* = 1, …, *L*}, thereby automating the significance testing process and preserving statistical power in various settings. Specifically, we propose to obtain a p-value via a *χ*^*2*^ test based on 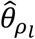 for the threshold *ρ*_*ℓ*_, 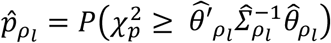, and construct a combined test statistic as: 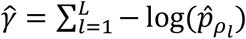 to calculate the final p-value for the testing of *H*_0_: *θ*_0_ = 0. For a given *ρ*_*l*_, we may obtain p-value 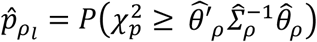 via a *p*-degree of freedom *χ*^*2*^ test, where 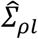 is the estimated variance-covariance matrix of 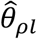 Since 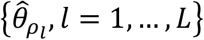 are estimated using overlapping data, the test statistics 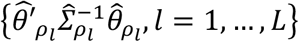 are highly correlated with each other. Thus, to estimate the null distribution of 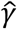, we use a resampling strategy to account for the correlations. Specifically, we generate a vector of *n*_*B*_ standard gaussian random variables 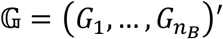, and subsequently obtain a perturbed random vector as 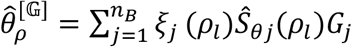, where

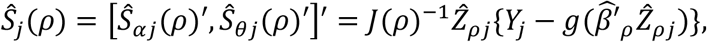

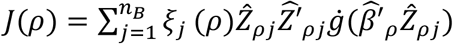 is the Fisher Information matrix for a given *ρ* and 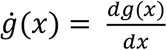. Subsequently, we obtain the perturbed counterpart of 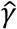 as 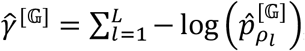, where

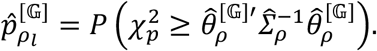

We then generate R realizations of 𝔾, {𝔾_*r*_, *r* = 1, …, *R*}, to obtain the final p-value for testing *H*_*0*_: *θ*_*0*_ = 0 as:

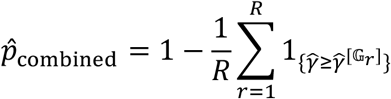

### 2.2. Data and metrics for evaluation

We evaluated the performance of ATLAS in simulation studies and conducted a real-world genetic association study using EHR data that has been linked to a biorepository.

#### 2.2.1. Simulation study

In our simulation studies, we estimated type I error rate (statistical size) as well as empirical statistical power of ATLAS under various settings using a publicly available de-identified database with *N* = 2,723 patient records containing 1,342 unique International Classification of Disease (ICD), Ninth Revision codes [15]. To test robustness of ATLAS under different linkage and inference settings, our simulations compared ATLAS performance by (1) creating discrepancies between databases by perturbing with multivariate noise, (2) adjusting the average codes per patient record, (3) adjusting effect sizes, and (4) using outputs from different linkage algorithms to test compatibility with ATLAS. We considered two linkage algorithms in our simulations: i) `ludic`, a published algorithm which relies on Bayesian modeling of binary diagnosis codes, and ii) `embeddingMatch`, which calculates cosine similarities between patient-level semantic embedding vectors (SEVs) similar to other embedding based linkage methods [15–18]. Further details regarding the `embeddingMatch` method and the simulation model are detailed in the Supplementary Appendix. Type I error rates using single cutoff thresholds and for the ATLAS threshold combination test were estimated as the proportion of p-values less than the nominal testing level *α* = 0.05 under no simulated association. Similarly, empirical power was estimated as the proportion of significant p-values at *α* = 0.05 under simulated association for a given OR > 1. Results are based on *n* = 1,500 simulations in each setting, and thresholds used in the ATLAS threshold combination test include *ρ* ∈ (0.1,0.3, …, 0.9). For the sake of simplicity, we report ATLAS results at single cuttoff thresholds of 0.1, 0.5, and 0.9, and report the rest in the Supplementary Appendix.

#### 2.2.2. Genetic association study using real-world biorepository data

To further validate performance and demonstrate real-world utility of ATLAS, we conducted a genetic association study with linked data to assess the association between rheumatoid arthritis (RA) genetic risk score (GRS) and various clinical outcomes among RA patients. We considered EHR records from two databases with which we performed linkage and the linked data-enabled downstream association analyses. The EHR data are stored in two separate databases: (i) the Crimson Clinical Discards database (herein referred to as Crimson and akin to database A) for a subset of RA patients of European descent previously identified in 2008 [19,20]; and (ii) the Mass General Brigham (MGB) Research Patient Data Registry (herein referred to as RPDR and akin to database B) subset of RA patients identified via an existing machine learning algorithm [21–23]. The Crimson RA cohort contains anonymized EHR data along with genotype data for *n*_*Crimson*_ = 1,284 patients collected up to 2008. The RPDR RA cohort contains full EHR data up to 2017 for a total of *n*_*RPDR*_ = 12,838 patients. A subset of *n*_*biobank*_ = 1,270 RPDR patients already report genotype data because they belong to the MGB Biobank, and we have gold standard labels on the true mappings between RPDR and MGB Biobank records. [24,25]. Our goal is to link Crimson with RPDR to enable genetic association studies of RA GRS against various clinical outcomes using all available data across RA cohorts. Figure 2 provides a schematic of our analyses. Details about how RA GRS were constructed are reported in the Supplementary Appendix [20,26,27].

**Figure 2:**
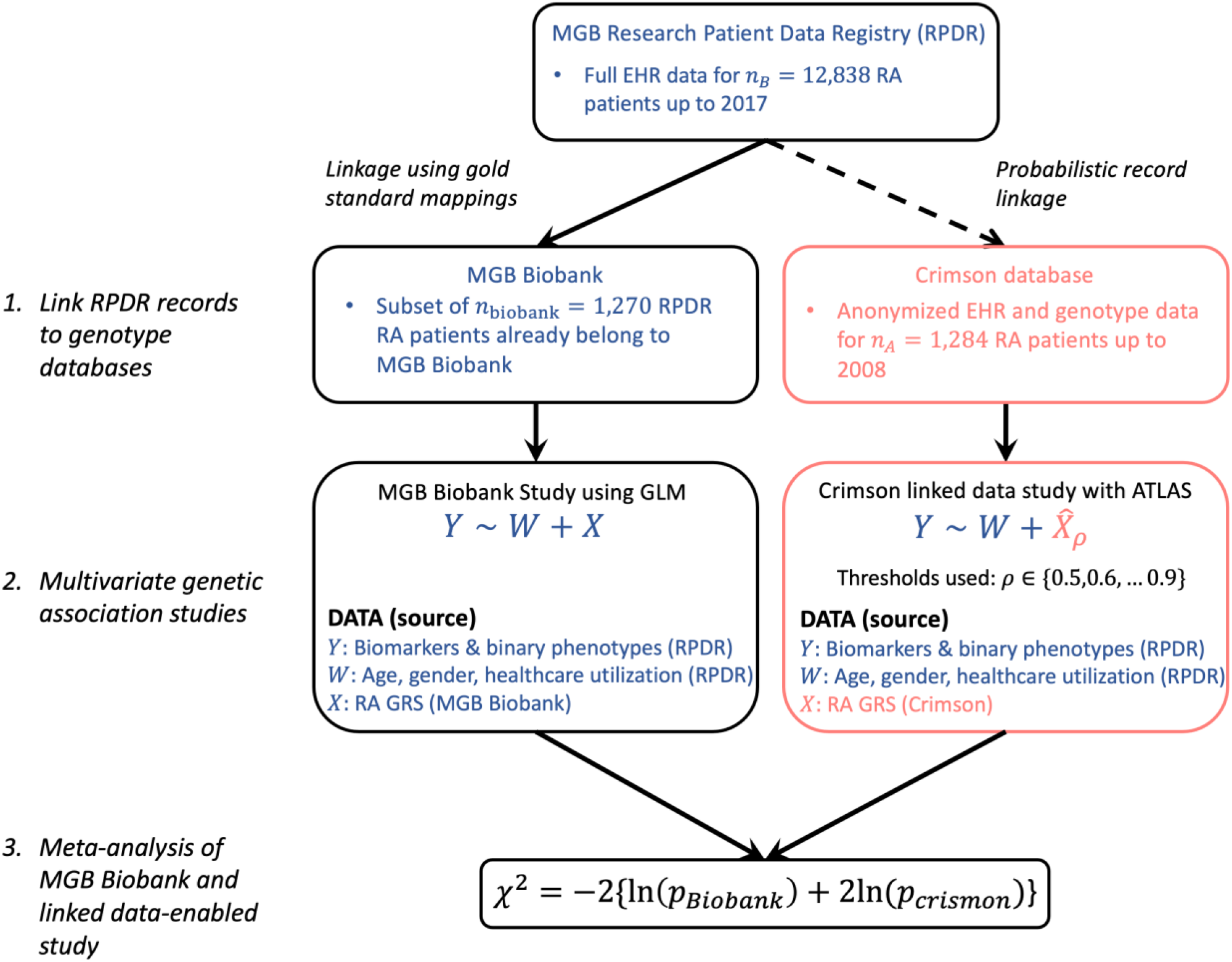
Schematic of real-world genetic association studies conducted using the MGB Biobank and the Crimson linked data.

To perform record linkage, we assembled all available ICD codes in Crimson records and ICD codes recorded prior and up to 2008 in RPDR records. ICD codes were then aggregated to PheCodes using the standard procedure, and a vector of 1,542 binary matching features was created for each patient record where each feature was a binary indicator of presence or abscence of a PheCode [28,29]. Next, we performed PRL using `ludic` to estimate probabilities of being a match for every possible pair of RPDR and Crimson records. Since the Crimson cohort consists of RA patients that were previously managed at MGB, we anticipated that a majority of these patients can be linked to the updated RPDR RA cohort and that some of the linked patients may report genotype data from both MGB Biobank and Crimson. In the absence of gold standard labels on the true mappings between RPDR and Crimson records, we validate the accuracy of the linkage by assessing the concordance between RA GRS from MGB Biobank and those from Crimson among the matched subset with genotype data available from both databases.

Once Crimson records are linked to RA RPDR records, we imputed GRS values using the weighted average method from Crimson to those subjects in RA RPDR using thresholds *ρ*_*ℓ*_ ∈ {0.5,0.6, …, 0.9}. Subsequently, we used ATLAS to conduct multivariate association studies of linked RA GRS and clinical outcomes while adjusting for patient age, gender, and healthcare utilization (defined as the log[1 + total encounters]). Clinical outcomes from RDPR include laboratory biomarkers commonly used to assess patient inflammation and binary phenotypes for pyogenic arthritis and gout, which are other distinct non-autoimmune forms of arthritis. These binary phenotypes were defined as having at least 2 PheCodes corresponding to these disorders and were constructed using ICD codes recorded up to 2017 in RPDR [28,29,29]. Reported effect sizes were estimated using data imputed at a *ρ* = 0.9 threshold.

Using RDPR patients who already belong to the MGB Biobank, we replicated these multivariate association studies and conducted meta-analysis using Fisher’s method to combine the P-values estimated from the MGB Biobank and Crimson linked-data study to demonstrate increased power to detect associations when linking databases. To determine statistical signifiance after meta-analysis, we accounted for multiple testing by adjusting p-values to control for a false discovery rate (FDR) of 5% using the Benjamini-Hochberg procedure [30].

#### 2.2.3. Benchmark methodology for comparison

In both our simulation studies and our real-world association study, we additionally considered the bias correcting estimators for linked data proposed by Han *et al*. as benchmark approaches [14]. The three Han *et al*. estimators are “Han F” (for “Full” – which considers all possible pairs for each patient in *A*), “Han M” (for “Max” – which considers largest probabilities for each patient in *A*), and “Han M2” (for “Max 2” – which considers two largest probabilities for each patient in *A*) [14]. We report type I error rates and empirical power of these estimators. We were unable to replicate the real-world multivariate study using Han *et al*. estimators because they do not consider accepting covariates from the database that provides outcomes. To compare performance in the real-world setting, we conducted univariate analyses.

## 3. Results

### 3.1. Type I error control in simulations

In Figure 2 we show estimated type I error rates of ATLAS under linkage conditions simulated according to (1) average number of codes per patient record and (2) discrepancies between the databases created by perturbing with multivariate noise. ATLAS effectively controlled for type I error in all simulation settings and at all considered thresholds regardless of the linkage or downstream imputation method used. Han et al.’s estimators controlled for type I error, although they appear somewhat too conservative.

### 3.2. Statistical power evaluation

Figure 3 shows empirical power of ATLAS using best match imputation. Under linkage conditions where patient records had on average 16 codes, there was little difference in power at different single-cutoff thresholds. Under conditions where patient records had on average 4 codes, we observed more significant differences in power between single cutoff thresholds. Relative power between thresholds was dependent on the linkage method. However, across all settings, power exhibited by the ATLAS threshold combination test demonstrated either the highest power or at least matched the highest power amongst ATLAS single cutoff thresholds. Larger simulated effect sizes increased power across all thresholds but did not change the relative power exhibited amongst thresholds. Empirical power when using weighted average imputation was similar in comparison to power using best match imputation, although differences in power based on imputation strategy were observed in certain settings when using embeddingMatch (Supplementary Figure 1). In comparison, the estimators proposed by Han *et al* did not capture signal as effectively as ATLAS in all simulation settings. Notably, when records had on average 4 codes, Han *et al*’s estimators failed to capture signal most likely due to noisy linkage probabilities generated from PRL.

**Figure 3:**
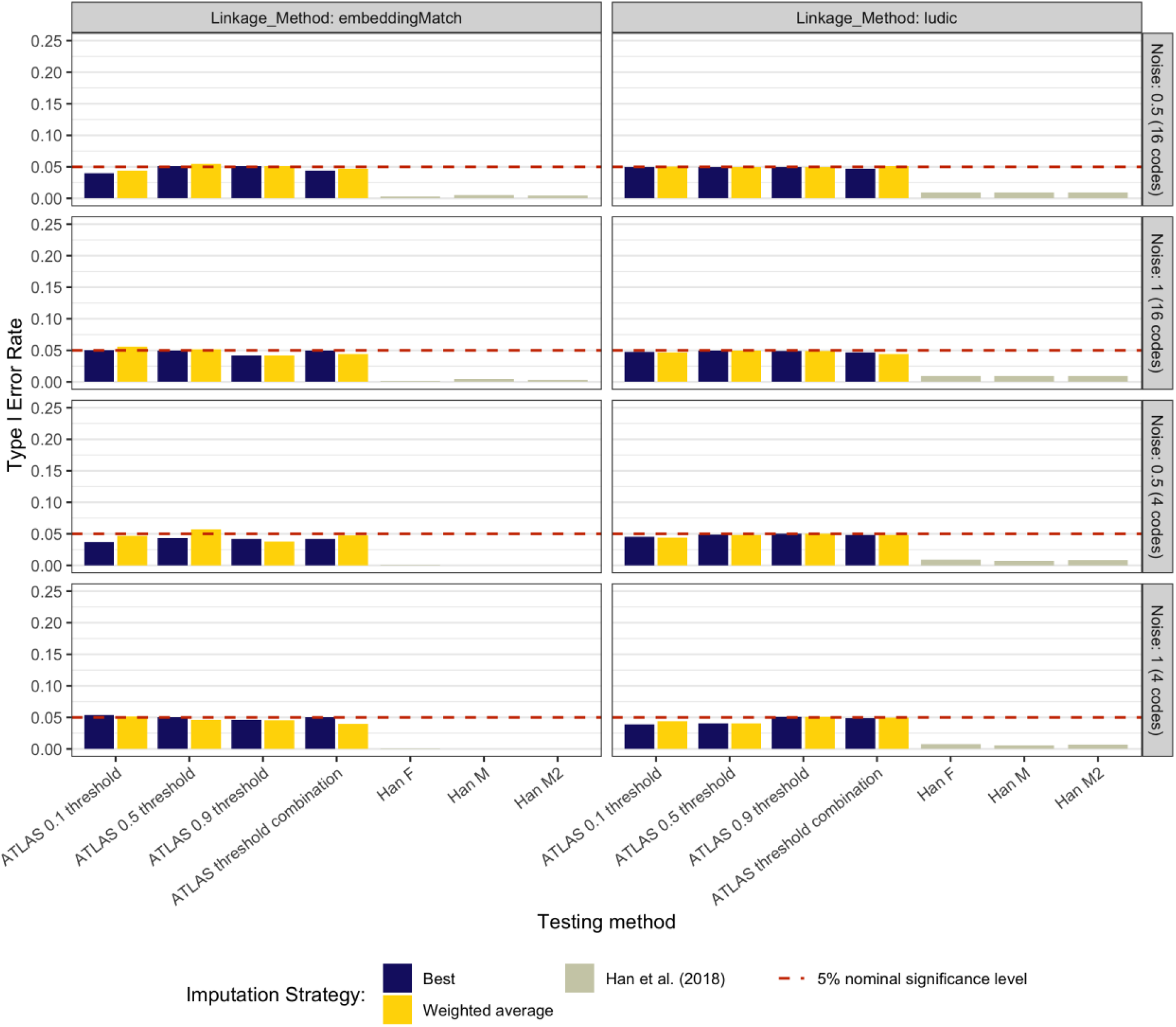
Comparison of type I error rates of ATLAS and Han et al. estimators in simulation settings with different noise levels and average codes per patient record (simulations under H_0_). ATLAS type I error rates reported for several single cutoff thresholds and the ATLAS threshold combination test.

We then evaluated ATLAS performance after creating false matches between databases to simulate linkage errors. In doing this, we sought to mimic rea l-world scenarios where linkage algorithms cannot discern between many pairs of patient records due to a high degree of overlapping features. ATLAS successfully controlled for Type I error in this setting (Supplementary Figure 2). In Figure 4, we show empirical power of ATLAS in presence of false matches. Most notably, use of the weighted average imputation yielded significantly higher power compared to best match imputed variables, and the ATLAS threshold combination test again demonstrated good power relative to its power from single cutoff thresholds. Han *et al*’s M2 estimator yielded comparable power in the context of false matches when using `ludic`, but was generally outperformed by the ATLAS threshold combination test.

**Figure 4:**
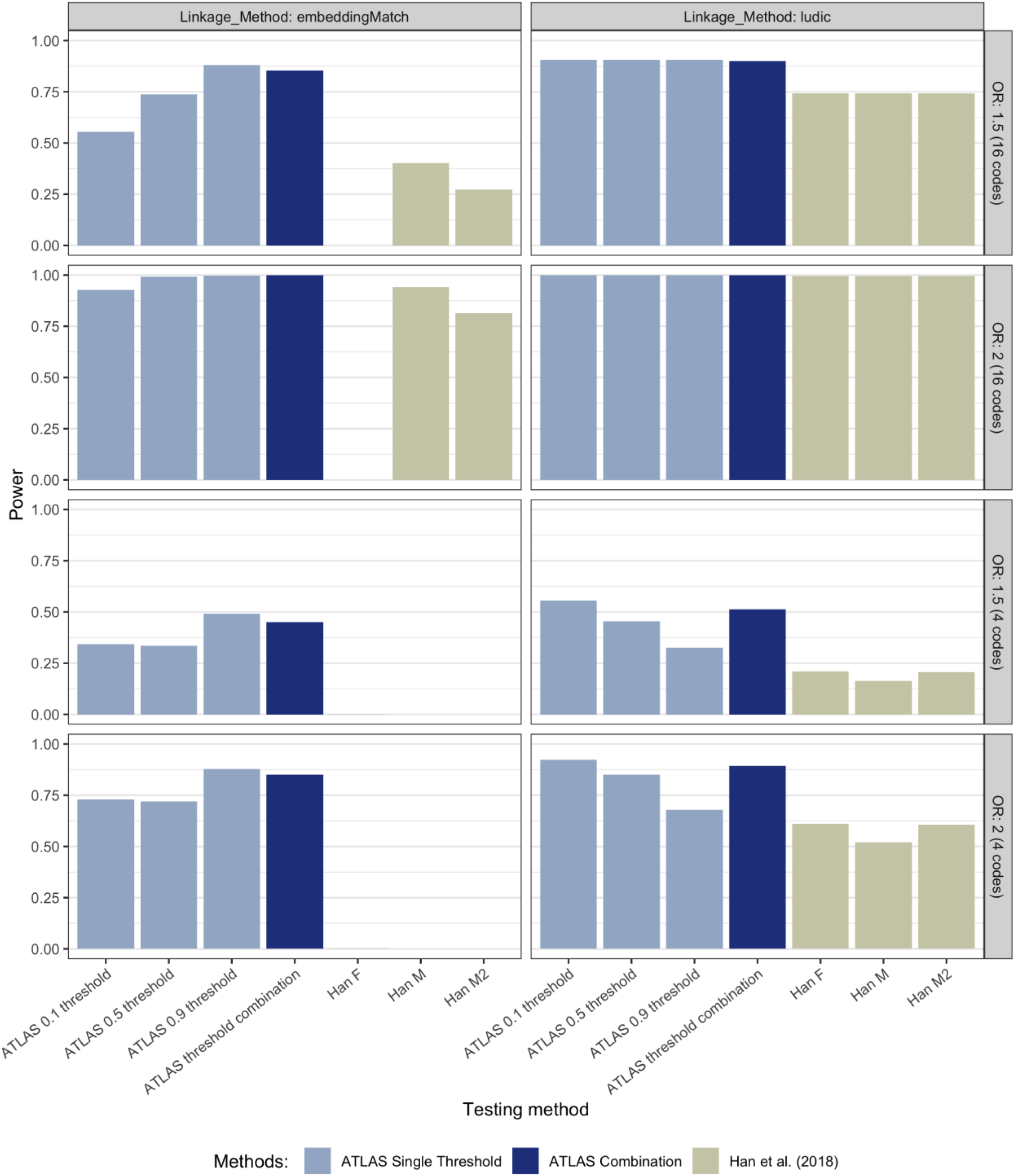
Comparison of empirical power of ATLAS and Han et al. estimators in simulation settings with different effect sizes and average codes per patient record (simulations under H_1_). Results were generated using the best match imputation method. ATLAS power reported for several single cutoff thresholds and the ATLAS threshold combination test.

### 3.3. Real-world genetics study: association of clinical outcomes with rheumatoid arthritis risk alleles among rheumatoid arthritis patients

We performed PRL on 12,838 patient records from RPDR and 1,284 patient records from Crimson. At a conservative threshold of *ρ* = 0.9, we identified 1,157 matching patient records between RPDR and Crimson. Out of 1,157 patients, we further identified 213 patients who have been genotyped in both RPDR and Crimson. Spearman’s correlation coefficient between the RPDR and Crimson GRS’ for these overlapping patients was estimated to be *ρ* = 0.83 (95% CI: 0.77-0.87, *P* = 2 *×* 10^*-16*^), suggesting high concordance of genetic data and reliable linkage quality. Univariate association study results using both ATLAS and Han *et al*. estimators are reported in Supplementary Table 1 and 2. As a positive control, we replicated the known association of anti-citrullinated protein antibody (ACPAs) levels with RA GRS using ATLAS and Han *et al*’s estimators [31,32]. In general, ATLAS detected larger effect sizes and smaller p-values compared to any of Han *et al*’s estimators, supporting simulation results that ATLAS is more powerful at detecting associations. For example, for log-transformed rheumatoid factor levels, ATLAS estimated *β*_GRS_ = 0.15 with *p* = 0.00 while Han *et al*’s M2 estimator estimated *β*_GRS_ = 0.14 with *p* = 0.09.

Multivariate association study results using the MGB Biobank and the Crimson linked data study are presented in Table 1 for biomarkers and Table 2 for phenotypes. Effect sizes estimated from both studies were generally concordant. Figure 5 visualizes the difference in adjusted p-values between using only the MGB Biobank cohort for which gold standard mappings were already available and after incorporating additional RA patients with genotype data through the Crimson linked cohort. We demonstrated improved power to detect associations when incorporating linked data as meta-analysis yielded two additional significant associations (namely log-transformed Erythrocyte sedimentation rate and C-Reactive Protein level). Further, the average unadjusted -log_10_(*P value*) among statistically significant outcomes was 6.45 in the MGB Biobank study and 9.65 after meta-analysis with the Crimson linked data study, again demonstrating the potential to increase power when incorporating additional data by linking databases.

**Table 1:** Multivariate association study results of RA GRS and patient biomarkers for the MGB Biobank RA cohort and the Crimson linked RA cohort. Crimson linked RA cohort effect sizes estimated a stringent imputation threshold of 0.9 and P-values estimated using the ATLAS combination test.

|  | **Beta** |  | **P-value** |  |  |
| --- | --- | --- | --- | --- | --- |
| Biomarker | MGB<br>Biobank (SE) | Crimson<br>linked—<br>ATLAS 0.9<br>threshold (SE) | MGB<br>Biobank | Crimson<br>linked—<br>ATLAS<br>combination | Two study P-value |
| Anti-citrullinated<br>protein antibodies<br>(log) | 0.39 (0.04) | 0.31 (0.04) | 9.39e-30 | 0.00e+00 | 8.54e-38 |
| Rheumatoid factor<br>(log) | 0.12 (0.03) | 0.16 (0.03) | 2.20e-06 | 0.00e+00 | 8.14e-15 |
| Erythrocyte<br>sedimentation rate | 0.49 (0.58) | 1.27 (0.55) | 3.91e-01 | 1.00e-02 | 2.56e-02 |
| C-reactive protein<br>(log) | 0.04 (0.02) | 0.04 (0.02) | 7.80e-02 | 3.00e-02 | 1.65e-02 |

**Table 2:** Multivariate association study results of RA GRS and binary phenotypes for the MGB Biobank RA cohort and the Crimson linked RA cohort. Crimson linked RA cohort effect sizes estimated a stringent imputation threshold of 0.9 and P-values estimated using the ATLAS combination test.

|  | **OR** |  | **P-value** |  |  |
| --- | --- | --- | --- | --- | --- |
| Phenotype | MGB<br>Biobank (SE) | Crimson linked—<br>ATLAS 0.9<br>threshold (SE) | MGB<br>Biobank | Crimson<br>linked—ATLAS<br>combination | Two study P-value |
| Pyogenic<br>arthritis | 1.41 (0.11) | 1.40 (0.18) | 1.64e-03 | 9.57e-02 | 1.12e-03 |
| Gout and other<br>crystal<br>arthropathies | 0.85 (0.07) | 0.85 (0.08) | 2.82e-02 | 2.43e-02 | 5.68e-03 |

**Figure 5:**
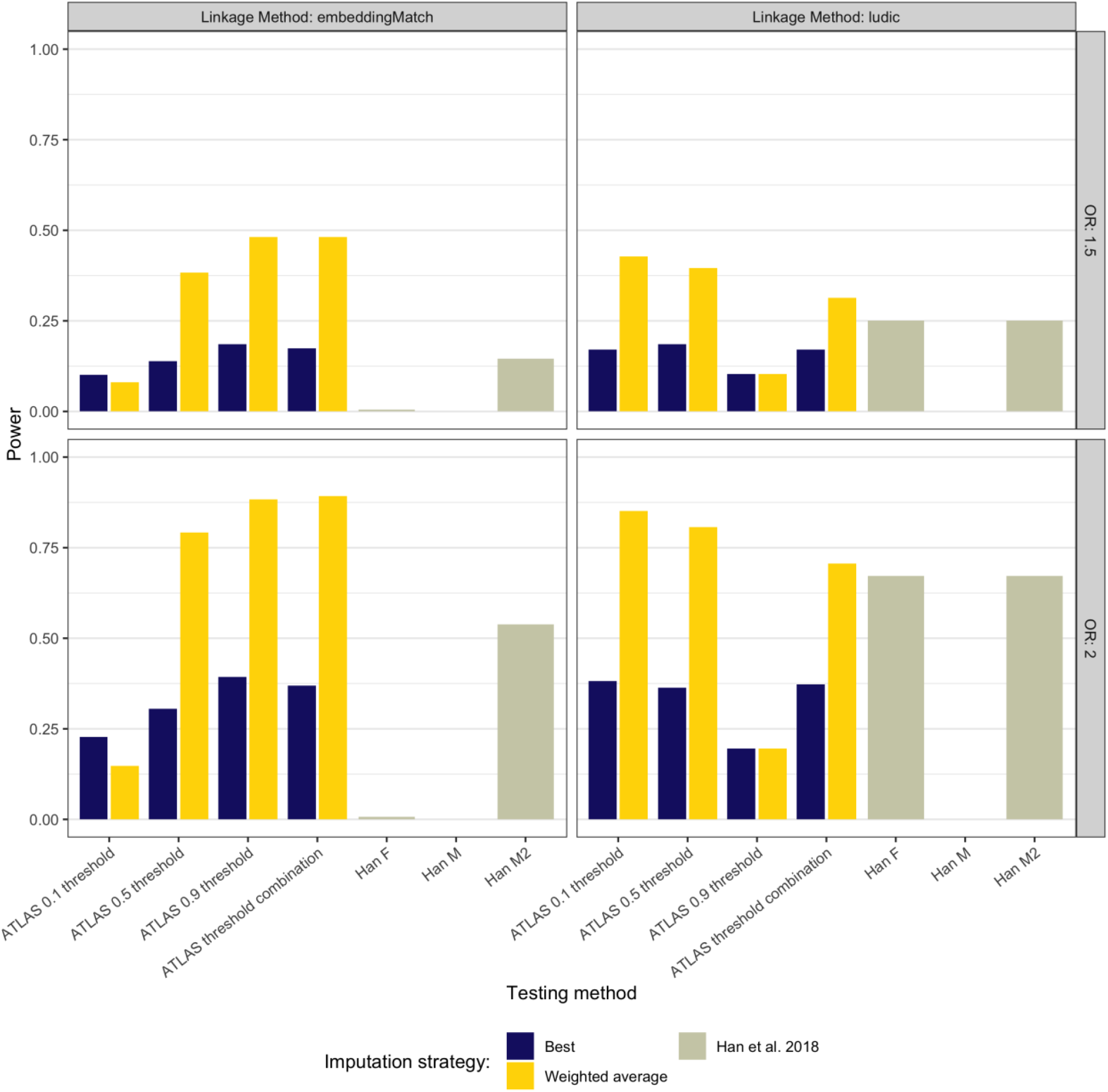
Comparison of empirical power of ATLAS and Han et al. estimators in presence of false matches between databases (simulations under H_1_). Simulated databases report on average 16 codes per patient record.

**Figure 6:**
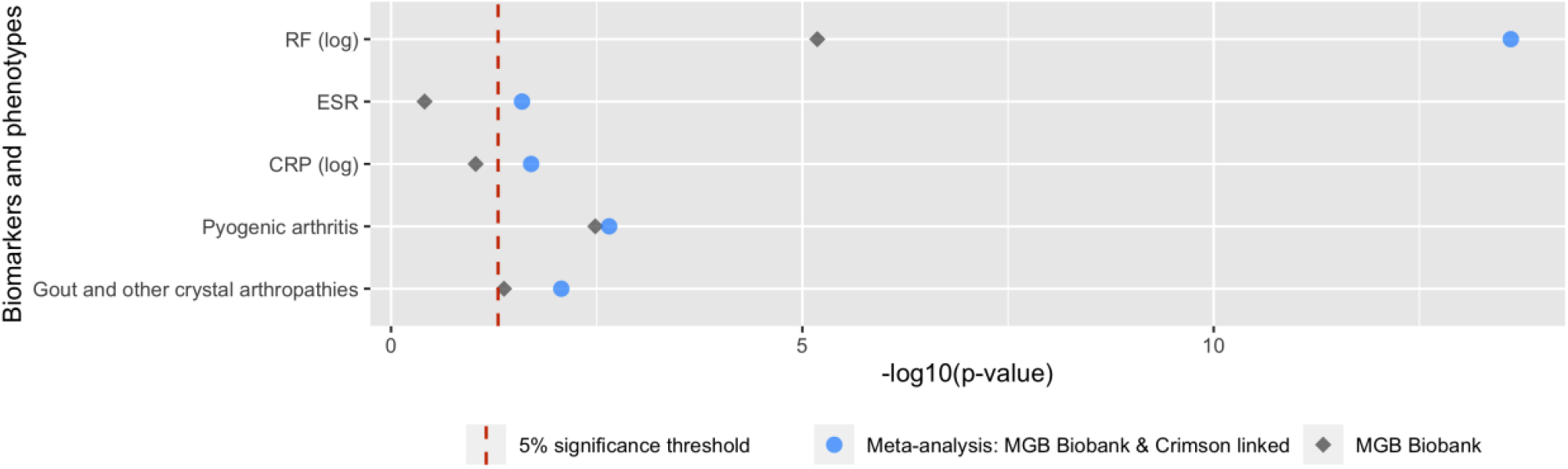
Logarithm transformed P-values from genetic association study using only RA patients with previously available genotype data at MGB Biobank and after incorporating additional RA patients with genotype data through the Crimson linked cohort.

## 4. Discussion

The tremendous amount of biomedical data becoming available for research has led to great interest and demand for linkage of databases, which allows researchers to capture more complete pictures of patient health and conduct novel research studies. Inference using linked data must acknowledge and mitigate bias in estimated effect sizes that are created by linkage errors while retaining good statistical power. To this end, we propose ATLAS as a supervised, robust, flexible, and scalable method that tests for association between variables originally belonging to separate databases. We demonstrate that ATLAS is a valid method that effectively controls for type I error regardless of linkage or imputation method used, and that ATLAS is more powerful than competing methods in a range of linkage and inference settings.

We demonstrate in simulation studies that weighted average imputation of missing variables not only protects against type I error in downstream inference but also preserves good statistical power to detect associations in the presence of linkage errors. Thus, it is the preferable imputation method to use in future studies to mitigate linkaged error induced bias. However, users should be aware that weighted average imputation with lower thresholds using linkage algorithms like `embeddingMatch` may yield lower power as seen in Supplementary Figure 1 when patient records contain few matching features.

During missing variable imputation, instead of selecting ad hoc thresholds above which patients are considered a match or naively imputing variables from pairs of patient records with the highest linkage probability like other proposed methods, ATLAS optimally combines p-values originating from various thresholds. Our results suggest three major advantages to the ATLAS threshold combination test: (1) avoids arbitrarily choosing a threshold for the linkage probability, thus completely automating the record linkage process; (2) reduces estimation bias by combining p-values estimated from data imputed at different thresholds; and (3) preserves good statistical power regardless of which threshold performs best in a given setting. Both simulation and application study results further demonstrate that the ATLAS threshold combination test substantially outperforms competing methods to detect associations in various settings. When selecting thresholds which are considered in the ATLAS threshold combination test, we caution against using every possible threshold in order to preserve good statistical power and recommend selecting no more than 10 thresholds.

These attractive features of ATLAS enable robust performance in real-world settings, and we demonstrated its utility in our real-world genetic study testing association between RA GRS and clinical outcomes. By meta-analyzing results obtained from MGB Biobank —for which genotype data was already available—and the newly linked Crimson cohort, we were able to increase power and detect two more associations than when using the MGB Biobank cohort alone. Our results suggest that higher GRS are associated with higher levels of inflammatory laboratory biomarkers, increased risk for pyogenic arthritis, and decreased risk for gout, This study demonstrates the potential to detect novel associations by expanding our sample size of patients at no additional cost and without further data collection efforts.

## 5. Conclusion

In this article, we introduce ATLAS, an automated and flexible algorithm that conducts robust inference using probabilistically linked databases. The ATLAS threshold combination te st exhibits high power to detect associations in a range of simulation settings while controlling for type I error, and it exhibits substantial improvement in power and flexibility over existing inference methods for linked databases. Thus, ATLAS promises to enable novel and powerful research studies using linked data.

## Supporting information

supplementary_information_ATLAS

## Data Availability

Datasets used in the simulation study are provided in the "ludic" package on CRAN.

## 7. Author Contributions

All authors made substantial contributions to: conception and design; acquisition, analysis, and interpretation of data; drafting the article or revising it critically for important intellectual content; and final approval of the version to be published.

## 8. Supplementary Material

Supplementary material is available at Journal of the American Medical Informatics Association online.

