## supplementary_information_ATLAS for "ATLAS: An automated association test using probabilistically linked health records with application to genetic studies"

### Supplementary Materials for ATLAS

#### 0. `embeddingMatch` Methodology

We used `embeddingMatch` as a linkage method in our simulations. Below, we provide details about this algorithm.

First, we constructed semantic embedding vectors (SEVs) for diagnosis codes (such as International Classification of Diseases — ICD — codes) by performing singular value decomposition (SVD) on the shifted positive pointwise mutual information (SPPMI) matrix as described in Beam *et al* [1,2]. For any given patient, we scanned through each of their codes as a target code. For any given target code occurring at time $t$, denoted by $w_t$ we counted all codes occuring within 30 days of $t$ as co-occurences with $w_t$. The total numbers of co-occurences for all possible pairs of codes are aggregated over all target codes within each patient and then across all patients, yielding the co-occurence matrix, denoted by $\mathbb{C} = [C(w,c)]$. Thus, given a target code $w$, we assume the context codes vocabulary $V_c(w)$ are the codes co-occurred with the target code within a 30-day window. We then calculated the SPPMI matrix as:

$$\text{SPPMI}(w,c) = \max\left\{\log\frac{C(w,c)}{C(w,\cdot)C(c,\cdot)} - \log(k), 0\right\}$$

with the negative sample $k$ set as 1 (i.e. no shifting), where $C(w,\cdot)$ is the row sum of $C(w,c)$ [2]. For each given SPPMI, we obtain its first $d$-dimensional SVD as $\mathbb{U}_d \text{diag}(\lambda_1, \ldots, \lambda_d)\mathbb{U}_d^T$ and then construct the $d$-dimensional embedding vectors as $\mathbb{V}_d$, where $\mathbb{V}_d = \mathbb{U}_d \text{diag}\sqrt{\lambda_1}, \ldots, \sqrt{\lambda_d}$.

Using trained code-level SEVs, we constructed patient-level SEVs as weighted sums of code-level SEVs to summarize the diagnosis codes contained within a patient record (using the standard weights created by sublinear term frequency-inverse document frequency — TF-IDF). Thus, for $k = 1, \ldots, K$ patient records and $j = 1, \ldots, J$ unique codes in a dataset, the $k^{th}$ patient-level SEV as can be defined and represented as:

$$SEV_k^{patient} = \sum_{j=1}^{J} SEV^j \cdot \log(1 + TF^{patient,j}) \cdot \log(\frac{N}{N_j})$$

where $TF^{patient,j}$ is the $j^{th}$ code frequency in patient record $k$, $N$ is total patient records, and $N_j$ is total patient records containing code $j$. For $s = 1, \ldots, S$ patient records in one dataset and $t = 1, \ldots, T$ patient records in the other, we computed the cosine similarity $\cos(s, t)$ between every pair of patient-level SEV in the two datasets, and disregarded any pairs yielding $\cos(s, t) < 0$. We then used these as pseudo probabilities for downstream association testing with ATLAS in simulations.

#### 1. Details of Simulation Set-up

To simulate probabilistic record linkage results, we used a publicly available de-identified database with $N = 2{,}723$ patient records containing 1,342 unique International Classification of Disease (ICD), Ninth Revision codes that we used as matching features [3]. Using the original database, we constructed different versions of database B by controlling for codes per patient record, [3]. Then, we simulated database A by modifying database B using correlated multivariate normal noise in each iteration of our simulations:

$$A_i^* = \mathbb{1}_{B_j u + (1 - B_j) v}$$

where $u \sim \mathcal{N}(1, \delta\Sigma)$, $v \sim \mathcal{N}(-3, \delta\Sigma)$ with $\Sigma$ the empirical correlation matrix of the original codes, B the original database, A the second altered database, $\delta$ the strength of altering noise introduced, and $\mathbb{1}_x$ the indicator function that equates to 1 if $x > 0$ and 0 otherwise. We then matched $n_B = 2{,}000$ patients from database B to $n_A = 1{,}523$ patients in database A with $n_T = 800$ true overlapping patients. Not all patients had a match, and since both databases are derived from the same original database, true matches were known.

We then simulated continuous allele counts and binary phenotypes for downstream inference. Simulated allele counts for database A were generated via binomial experiments with a size parameter equal to 2 and a probability of success corresponding to the minor allele frequency (MAF), which we set to be $\text{MAF} = 0.2$. Simulated binary phenotype $Y$ for database B were generated from a logistic regression model:

$$\text{logit}(Y) = -0.5 + \log(\text{OR})X,$$

where $OR$ is the set odds ratio and $X$ is the allele count.

#### 2. Rheumatoid Arthritis Genetic Risk Score Composition

To construct rheumatoid arthritis (RA) genetic risk scores (GRS), we utilized whole-genome genotype data to construct human leukocyte antigen (HLA) and non-HLA GRS. We then summed each patient’s HLA and non-HLA GRS to calculate a composite RA GRS. Amino acid polymorphisms in the HLA region of chromosome 6 that have an established association with RA disease were used to calculated HLA GRS [4]. Single nucleotide polymorphisms (SNPs) outside of the HLA region that were previously shown to be significantly associated with RA in genome-wide association studies (GWAS) were used to calculate non-HLA GRS [5].

#### 2.1. HLA GRS

SNP2HLA was first used to impute amino acid polymorphisms in the HLA region, and the specific amino acid combinations on the HLA-DRB1 gene that Raychaudhuri *et al.* established to be significantly associated with RA were identified [4,6]. HLA GRS were constructed as

$$GRS_{HLA} = \sum_{i=1}^{p} w_i X_i$$

, where $p$ is the number of considered amino acid residue combinations, $w_i$ is the assigned weight for each amino acid combination, and $X_i$ is a binary indicator of the presence or absence of specific amino acid combinations considered.

#### 2.2. Non-HLA GRS

For $n$ non-human leukocyte antigen (HLA) SNPs,

$$GRS_{non-HLA} = \sum_{i=1}^{n} w_i Y_i$$

, where $n$ is the number of SNPs, $w_i$ is the assigned weight for the SNP, and $Y_i$ is the number of risk alleles (0, 1, or 2) [7]. SNP weights were derived from a published genome-wide association study (GWAS) meta-analysis, and non-HLA SNPs used to construct RA GRS are reported in Table S3 [5].

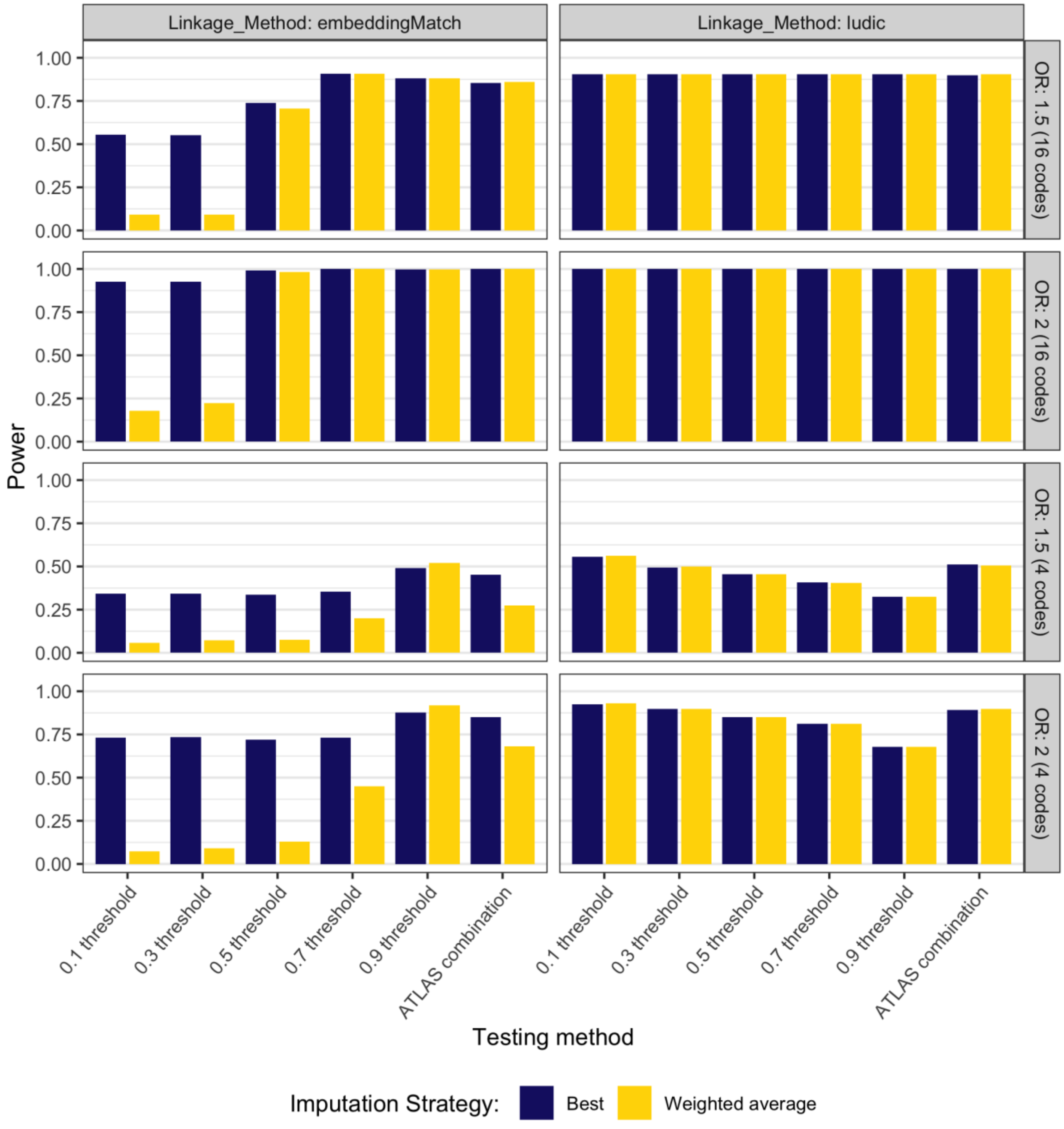


*Figure S1: Comparison of ATLAS empirical power at different single cutoff thresholds and the ATLAS threshold combination test in simulation settings with different effect sizes and average codes per patient record (simulations under $H_1$).*

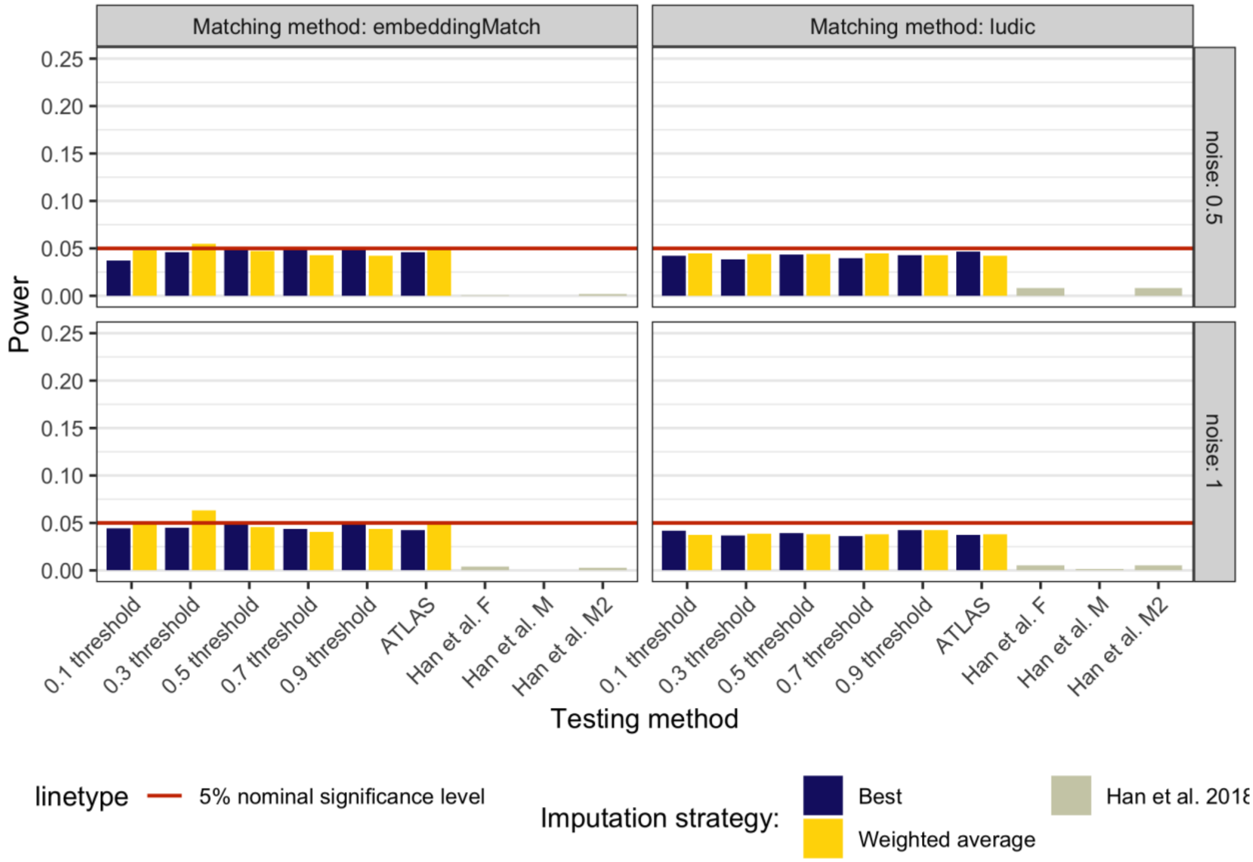


*Figure S2: Comparison of type I error rates in the presence of false matches in simulation settings with different noise levels and average codes per patient record (simulations under $H_0$).*

| | Beta | | | | | P-value | | | | |
|---|---|---|---|---|---|---|---|---|---|---|
| | MGB Biobank | Crimson linked | | | | MGB Biobank | Crimson linked | | | |
| Biomarker | | ATLAS 0.9 threshold (SE) | Han F (SE) | Han M (SE) | Han M2 (SE) | | ATLAS combination test | Han F | Han M | Han M2 |
| Anti-citrullinated protein antibodies (log) | 0.39 (0.03) | 0.31 (0.04) | 0.31 (0.06) | 0.30 (0.07) | 0.31 (0.07) | 0.00e+0.00 | 0.00e+00 | 9.55e-0.8 | 4.69e-05 | 1.57e-06 |
| Rheumatoid factor (log) | 0.12 (0.03) | 0.15 (0.03) | 0.13 (0.06) | 0.13 (0.07) | 0.14 (0.07) | 5.99e-06 | 0.00e+00 | 1.01e-01 | 1.56e-01 | 9.25e-02 |
| Erythrocyte sedimentation rate | 0.00 (0.63) | 0.72 (0.62) | 0.55 (0.69) | 0.65 (0.99) | 0.68 (0.69) | 9.98e-01 | 1.76e-01 | 4.30e-01 | 5.11e-01 | 3.22e-01 |
| C-reactive protein (log) | 0.03 (0.03) | 0.03 (0.02) | 0.00 (0.03) | 0.00 (0.04) | 0.01 (0.03) | 2.59e-01 | 7.14e-02 | 9.56e-01 | 9.46e-01 | 8.20e-01 |

*Table S1: Univariate association study results of RA GRS and patient biomarkers for the MGB Biobank RA cohort and the Crimson linked RA cohort. ATLAS estimated effect sizes using the Crimson linked RA cohort estimated using a stringent imputation threshold of 0.9 and P-values estimated using the ATLAS combination test.*

| | OR | | | | | P-value | | | | |
|---|---|---|---|---|---|---|---|---|---|---|
| | MGB Biobank | Crimson linked | | | | MGB Biobank | Crimson linked | | | |
| Phenotype | | ATLAS 0.9 threshold (SE) | Han F (SE) | Han M (SE) | Han M2 (SE) | | ATLAS combination test | Han F | Han M | Han M2 |
| Gout and other crystal arthropathies | 0.82 (0.07) | 0.83 (0.07) | 0.80 (0.06) | 0.83 (0.06) | 0.82 (0.05) | 3.15e-03 | 1.29e-02 | 3.30e-04 | 1.11e-03 | 3.69e-04 |
| Pyogenic arthritis | 1.31 (0.10) | 1.19 (0.09) | 1.23 (0.13) | 1.20 (0.11) | 1.21 (0.11) | 9.00e-03 | 8.57e-02 | 1.17e-01 | 9.14e-02 | 9.11e-02 |

*Table S2: Univariate association study results of RA GRS and binary phenotypes for the MGB Biobank RA cohort and the Crimson linked RA cohort. ATLAS estimated effect sizes using the Crimson linked RA cohort estimated using a stringent imputation threshold of 0.9 and P-values estimated using the ATLAS combination test.*

*Table S3: Non-HLA RA risk SNPS as identified in Okada et al. used to construct non-HLA GRS*

| SNP | Chromosome | Base Pair | Gene |
|---|---|---|---|
| rs227163 | 1 | 7,961,206 | TNFRSF9 |
| rs2301888 | 1 | 17,672,730 | PADI4 |
| rs28411352 | 1 | 38,278,579 | MTF1-INPP5B |
| rs12140275 | 1 | 38,633,879 | LOC339442 |
| rs2476601 | 1 | 114,377,568 | PTPN22 |
| rs624988 | 1 | 117,263,790 | CD2 |
| rs2228145 | 1 | 154,426,970 | IL6R |
| rs2317230 | 1 | 157,674,997 | FCRL3 |
| rs4656942 | 1 | 160,831,048 | LY9-CD244 |
| rs72717009 | 1 | 161,405,053 | FCGR2A |
| rs2105325 | 1 | 173,349,725 | LOC100506023 |
| rs17668708 | 1 | 198,640,488 | PTPRC |
| rs10175798 | 2 | 30,449,594 | LBH |
| rs34695944 | 2 | 61,124,850 | REL |
| rs13385025 | 2 | 62,461,120 | B3GNT2 |
| rs1858037 | 2 | 65,598,300 | SPRED2 |
| rs9653442 | 2 | 100,825,367 | AFF3 |
| rs6732565 | 2 | 111,607,832 | ACOXL |
| rs11889341 | 2 | 191,943,742 | STAT4 |
| rs6715284 | 2 | 202,154,397 | CFLAR-CASP8 |
| rs1980422 | 2 | 204,610,396 | CD28 |
| rs3087243 | 2 | 204,738,919 | CTLA4 |
| rs4452313 | 3 | 17,047,032 | PLCL2 |
| rs3806624 | 3 | 27,764,623 | EOMES |
| rs73081554 | 3 | 58,302,935 | DNASE1L3-ABHD6-PXK |
| rs9826828 | 3 | 136,402,060 | IL20RB |
| rs11933540 | 4 | 26,120,001 | C4orf52 |
| rs2664035 | 4 | 48,220,839 | TEC |
| rs45475795 | 4 | 123,399,491 | IL2-IL21 |
| rs7731626 | 5 | 55,444,683 | ANKRD55 |
| rs2561477 | 5 | 102,608,924 | C5orf30 |
| rs657075 | 5 | 131,430,118 | IL3-CSF2 |
| chr6:14103212 | 6 | 14,103,212 | CD83 |
| rs9268839 | 6 | 32,428,772 | HLA-DRB1 |
| rs2234067 | 6 | 36,355,654 | ETV7 |

| | | | |
|---|---|---|---|
| rs9372120 | 6 | 106,667,535 | ATG5 |
| rs17264332 | 6 | 138,005,515 | TNFAIP3 |
| rs7752903 | 6 | 138,227,364 | TNFAIP3 |
| rs2451258 | 6 | 159,506,600 | TAGAP |
| rs1571878 | 6 | 167,540,842 | CCR6 |
| rs67250450 | 7 | 28,174,986 | JAZF1 |
| rs4272 | 7 | 92,236,829 | CDK6 |
| chr7:128580042 | 7 | 128,580,042 | IRF5 |
| rs2736337 | 8 | 11,341,880 | BLK |
| rs998731 | 8 | 81,095,395 | TPD52 |
| rs678347 | 8 | 102,463,602 | GRHL2 |
| rs1516971 | 8 | 129,542,100 | PVT1 |
| rs11574914 | 9 | 34,710,338 | CCL19-CCL21 |
| rs10985070 | 9 | 123,636,121 | TRAF1-C5 |
| rs706778 | 10 | 6,098,949 | IL2RA |
| rs947474 | 10 | 6,390,450 | PRKCQ |
| rs3824660 | 10 | 8,104,722 | GATA3 |
| rs12413578 | 10 | 9,049,253 | 10p14 |
| rs793108 | 10 | 31,415,106 | ZNF438 |
| rs2671692 | 10 | 50,097,819 | WDFY4 |
| rs71508903 | 10 | 63,779,871 | ARID5B |
| rs726288 | 10 | 81,706,973 | SFTPD |
| rs331463 | 11 | 36,501,787 | TRAF6-RAG1/2 |
| rs508970 | 11 | 60,906,450 | CD5 |
| rs968567 | 11 | 61,595,564 | FADS1-FADS2-FADS3 |
| rs11605042 | 11 | 72,411,664 | ARAP1 |
| rs4409785 | 11 | 95,311,422 | CEP57 |
| rs10790268 | 11 | 118,729,391 | CXCR5 |
| rs73013527 | 11 | 128,496,952 | ETS1 |
| rs773125 | 12 | 56,394,954 | CDK2 |
| rs1633360 | 12 | 58,108,052 | CDK4 |
| rs10774624 | 12 | 111,833,788 | SH2B3-PTPN11 |
| rs9603616 | 13 | 40,368,069 | COG6 |
| rs3783782 | 14 | 61,940,675 | PRKCH |
| rs1950897 | 14 | 68,760,141 | RAD51B |
| rs8032939 | 15 | 38,834,033 | RASGRP1 |
| rs8026898 | 15 | 69,991,417 | LOC145837 |

| | | | |
|---|---|---|---|
| rs4780401 | 16 | 11,839,326 | TXNDC11 |
| rs13330176 | 16 | 86,019,087 | IRF8 |
| rs72634030 | 17 | 5,272,580 | C1QBP |
| rs1877030 | 17 | 37,740,161 | MED1 |
| chr17:38031857 | 17 | 38,031,857 | IKZF3-CSF3 |
| rs8083786 | 18 | 12,881,361 | PTPN2 |
| rs2469434 | 18 | 67,544,046 | CD226 |
| rs34536443 | 19 | 10,463,118 | TYK2 |
| chr19:10771941 | 19 | 10,771,941 | ILF3 |
| rs4239702 | 20 | 44,749,251 | CD40 |
| rs73194058 | 21 | 34,764,288 | IFNGR2 |
| chr21:35928240 | 21 | 35,928,240 | RCAN1 |
| rs8133843 | 21 | 36,738,242 | RUNX1-LOC100506403 |
| rs1893592 | 21 | 43,855,067 | UBASH3A |
| rs11089637 | 22 | 21,979,096 | UBE2L3-YDJC |
| rs3218251 | 22 | 37,545,505 | IL2RB |
| rs909685 | 22 | 39,747,671 | SYNGR1 |
